# COVID-19 (Coronavirus Disease) Outbreak Prediction Using a Susceptible-Exposed-Symptomatic Infected-Recovered-Super Spreaders-Asymptomatic Infected-Deceased-Critical (SEIR-PADC) Dynamic Model

**DOI:** 10.1101/2020.12.20.20240507

**Authors:** Ahmad Sedaghat, Amir Mosavi

## Abstract

Extension of SIR type models has been reported in a number of publications in mathematics community. But little is done on validation of these models to fit adequately with multiple clinical data of an infectious disease. In this paper, we introduce SEIR-PAD model to assess susceptible, exposed, infected, recovered, super-spreader, asymptomatic infected, and deceased populations. SEIR-PAD model consists of 7-set of ordinary differential equations with 8 unknown coefficients which are solved numerically in MATLAB using an optimization algorithm to fit 4-set of COVID-19 clinical data consist of cumulative populations of infected, deceased, recovered, and susceptible. Trends of COVID-19 in Trends in Gulf Cooperation Council (GCC) countries are successfully predicted using available data from outbreak until 23rd June 2020. Promising results of SEIR-PAD model provide insight into better management of COVID-19 pandemic in GCC countries.

## I. Introduction

Epidemiological models had shown to be essential in modeling, analysis and projection of the pandemics. In 2012, the Middle Eastern respiratory syndrome coronavirus (MERS-CoV) was reported with largest impact in Saudi Arabia, United Arab Emirates, and South Korea which caused more than 35% mortality rate among infected people with fever, cough, and shortness of breath [1, 2]. This raised concerns in the largest gathering event (Hajj) due to super-spreaders population from the locals infected by MERS-CoV. Alasmawi et al. [3] studied MERS-CoV by an extended SIR model with 5 set of ordinary differential equations for two main subcategories: local population and non-local pilgrims. The 5 set of population include susceptible (S), infected (I), super-spreaders (P), recovered (R), and hospitalized (H). Their study focused on finding mathematically the re-production number (R0) and they model coefficients merely found from literature. Kim et al. [4] studied MERS-CoV which caused the major outbreak in South Korea in 2015. They used a similar extended SIR model as [3] except with 6 set of ordinary differential equations adding asymptomatic infected population (A) with emphasis in determining transmission rates using estimation or literature. Only one set of clinical data on cumulative incidence numbers were used for model validation suggesting that the super spreader population caused initial large reproduction number. They concluded preventive measures such as intensive lockdown and quarantine hospitalized cases had major impact on stopping spread of the MERS-CoV outbreak.

Ndaïrou et al. [5] have recently applied the same extended SIR type model introduced in [3, 4] for studying outbreak of COVID-19 in Wuhan, China with emphasis in stability of their model with 8 set of ordinary differential equations including mortality rates. They fitted their model with only two set of clinical data including confirmed daily cases and confirmed daily death cases in Wuhan, China. They estimated model coefficients with values so that a reproduction number below the value of one is obtained. The SIR type models are usually designed for fitting with cumulative clinical data than daily data.

More recently, Xue et al. [6] developed a 7 set of equations SIR type model using a transmission network to fit COVID-19 clinical data in Wuhan (China), Toronto (Canada), and the Italy. They used an optimization algorithm (MCMC) to fit coefficients of 7 set of ODE equations. They compare only their model with two set of clinical data, i.e. confirmed total infected cases and confirmed active infected cases.

Observing the above studied literature, it is not clear why 5 to 8 set of ordinary differential equations are used to only fit with one or maximum two clinical data. From recent COVID-19 data, it can be easily seen that at least 5 set of different clinical data including susceptible, infected, recovery, critical and deceased populations can be used for validation of extended SIR type models. In this paper, we introduce SEIR-PADC which is SIR type model with 8 set of ODE equations. The aim of the paper is to computationally study the 5 set of clinical data on COVID-19 in GCC countries [7] simultaneously to be fitted by SEIR-PADC model using optimization technique in MATLAB (fminsearch tool) and measure clinical significance on development of COVID-19 in GCC countries.

## II. Materials and methdos

### A. SEIR-PADC Model

A We introduce here SEIR-PADC model consist of susceptible (S), exposed (E), symptomatic infected (I), recovered (R), super spreaders (P), asymptomatic infected (A), deceased (D), and critical (C) populations. The model consist of 8 different cumulative populations and transmission from each population starts similar to original SIS and SIR model reported by Kermack andMcKendrick [8]. A flow chart of SEIR-PADC model is shown in Figure 1.

**Figure 1:**
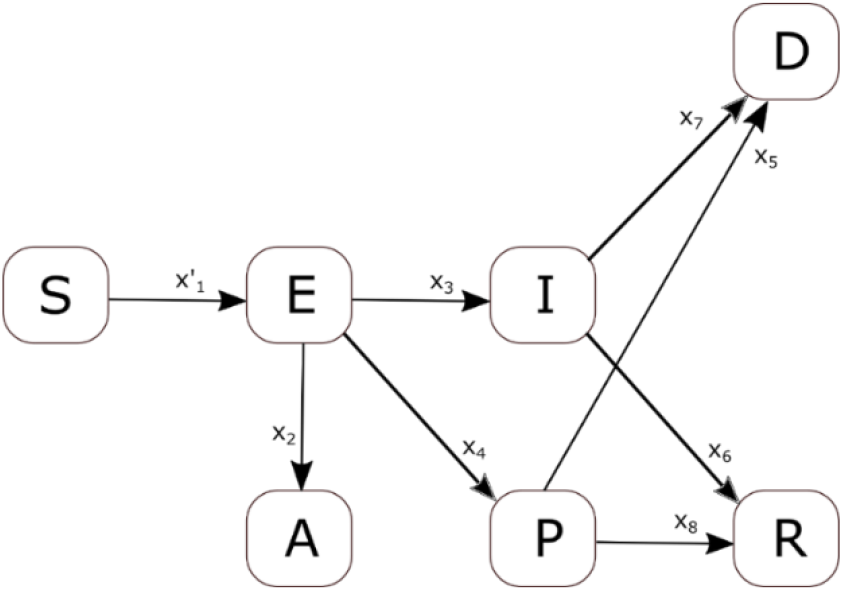
Flow chart of SEIR-PAD dynamic model.

The model have 12 unknown transmission coefficients x=[x_1_ x_2_ …x_12_], which all are positive or negative constants and are obtained by an optimization algorithm to fit clinical data. In Figure 1, x’_1_=x_1_E/N is selected as introduced in [8] and flow in and out of each population is simply expressed as follows.

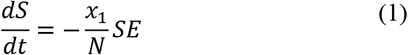

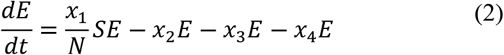

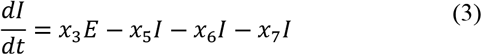

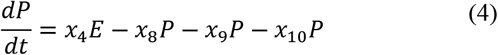

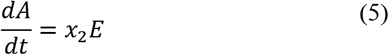

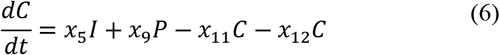

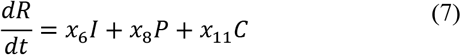

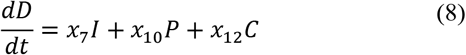

The total population *N* is constant and is defined by:

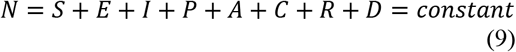

Kuwait has a population around 4,270,571 based on Worldometer 2020 [9]. A set of initial conditions are applied for the 8 populations and 12 coefficients to solve the set of 8 ODE equations (1-8) as follows.

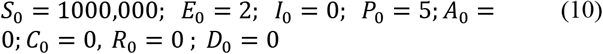

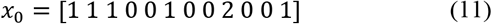

The SEIR-PADC model outlined above are solved using ode45 in MATLAB [10]. The optimization tool (fminsearch) [11] is used to find best fitted ODE coefficients by applying the following convergence criteria for 5-set of available data on goodness of fit (GOF).

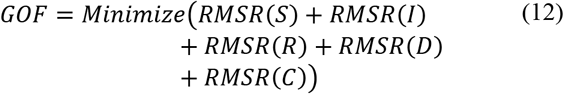

In equation (12), the root mean-square ratio (RMSR) of computed population versus available COVID-19 population are calculated and the optimization algorithm (fminsearch) in MATLAB is used to minimize the summation. This is discussed next.

### B. Goodness of fit technique; the root mean-square ratio

The root mean-square ratio (RMSR) is defined here using the coefficient of determination which is widely used on goodness of fit when predicted variables are compared with actual data particularly for situations that future outcomes are sought. The coefficient of determination (R^2^) evaluates the predicted values (*y*) against actual data (*y*_*exp*_) and is rewritten to provide RMSR as follows [12,13].

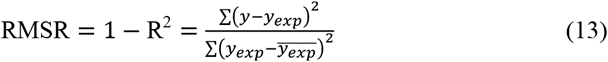

In equation (13), 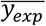 is the average of COVID-19 clinical data value for certain population. Better fitted functions provide R^2^ value close to unity and RMSR close to zero.

## III. Results

### A. SEIR-PADC model

Figure 2 shows that 5 set of COVID-19 data including critical, infected, deceased, recovered, and susceptible populations are simultaneously fitted with SEIR-PADC model. Goodness of fit value (GOF) is obtained 1.5756 for 5-set of data in equation (12).

**Figure 2:**
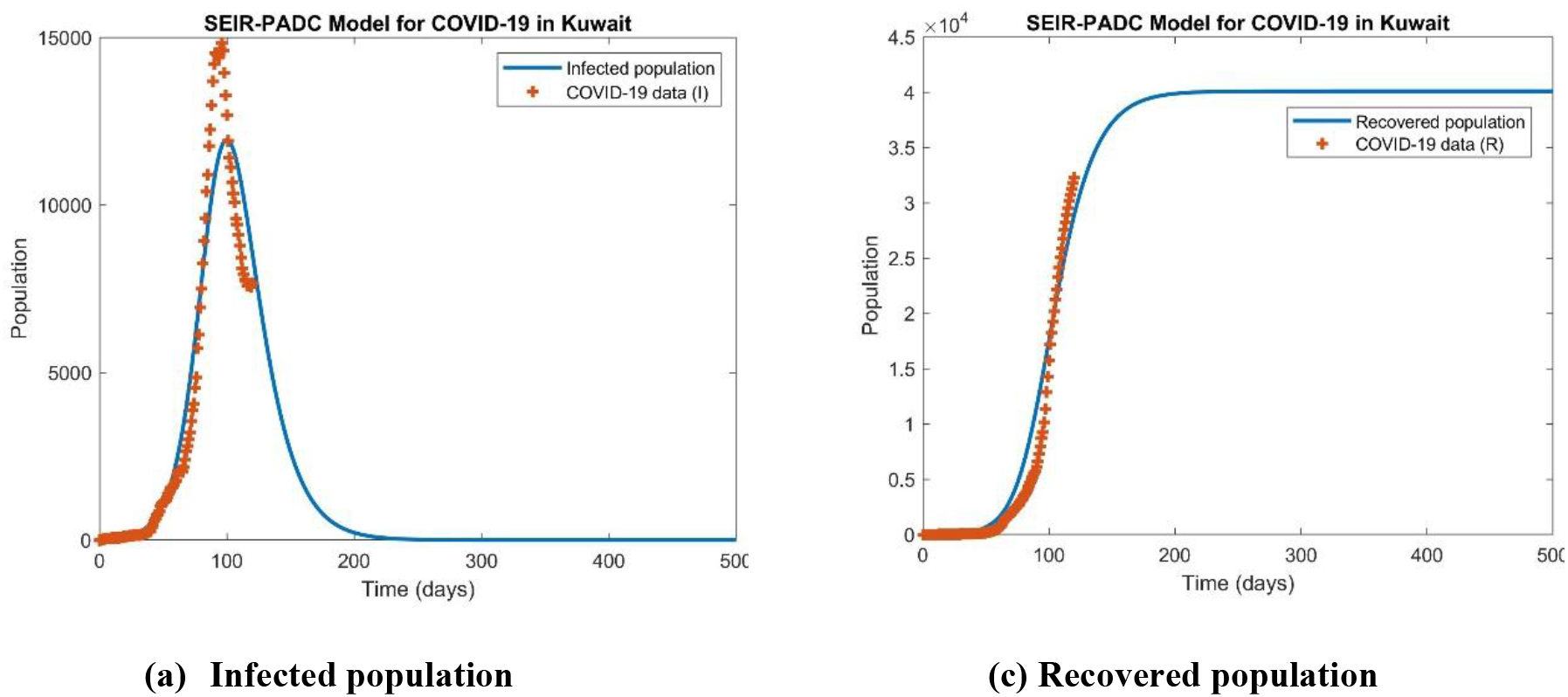
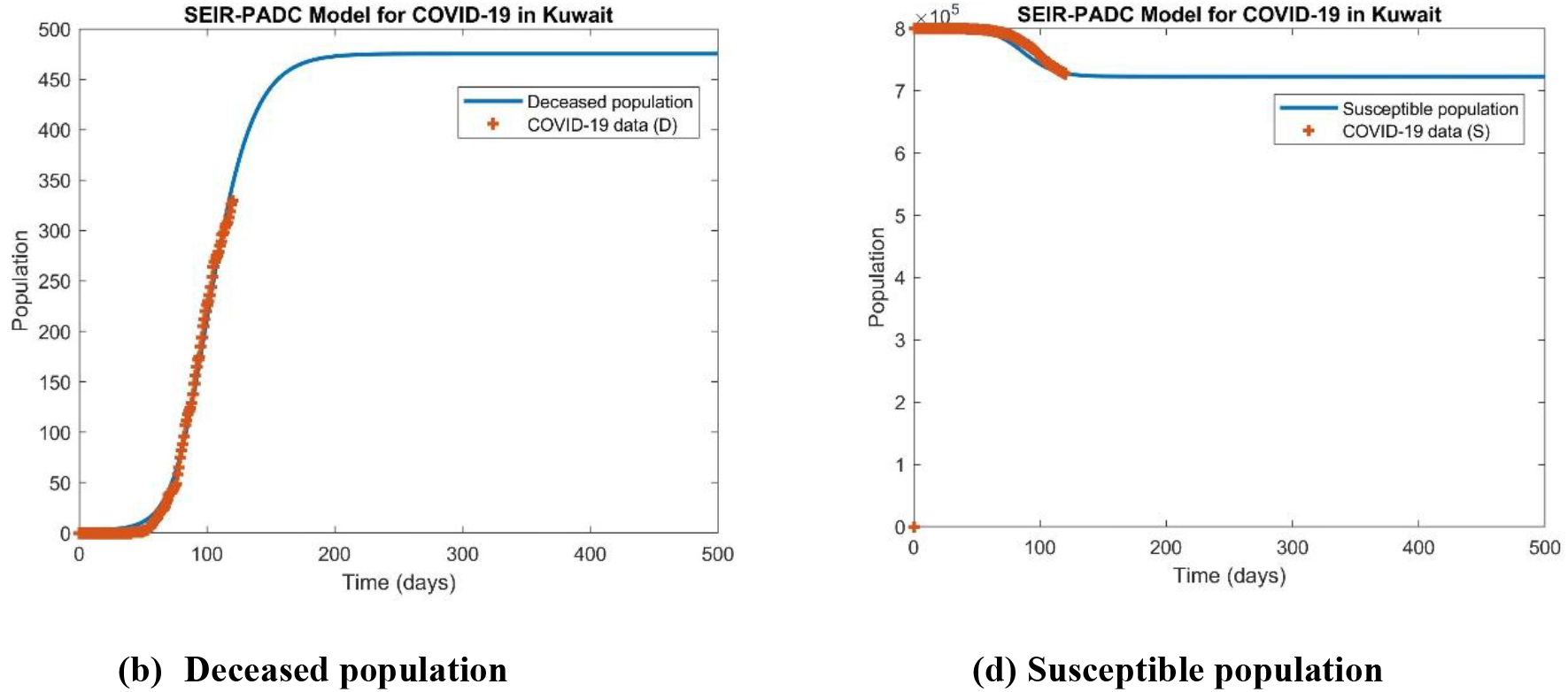
Fitting 4 set of COVID-19 data simultaneously with SEIR-PADC model for dynamics of COVID-19 in Kuwait (23 June 2020).

**Figure 3:**
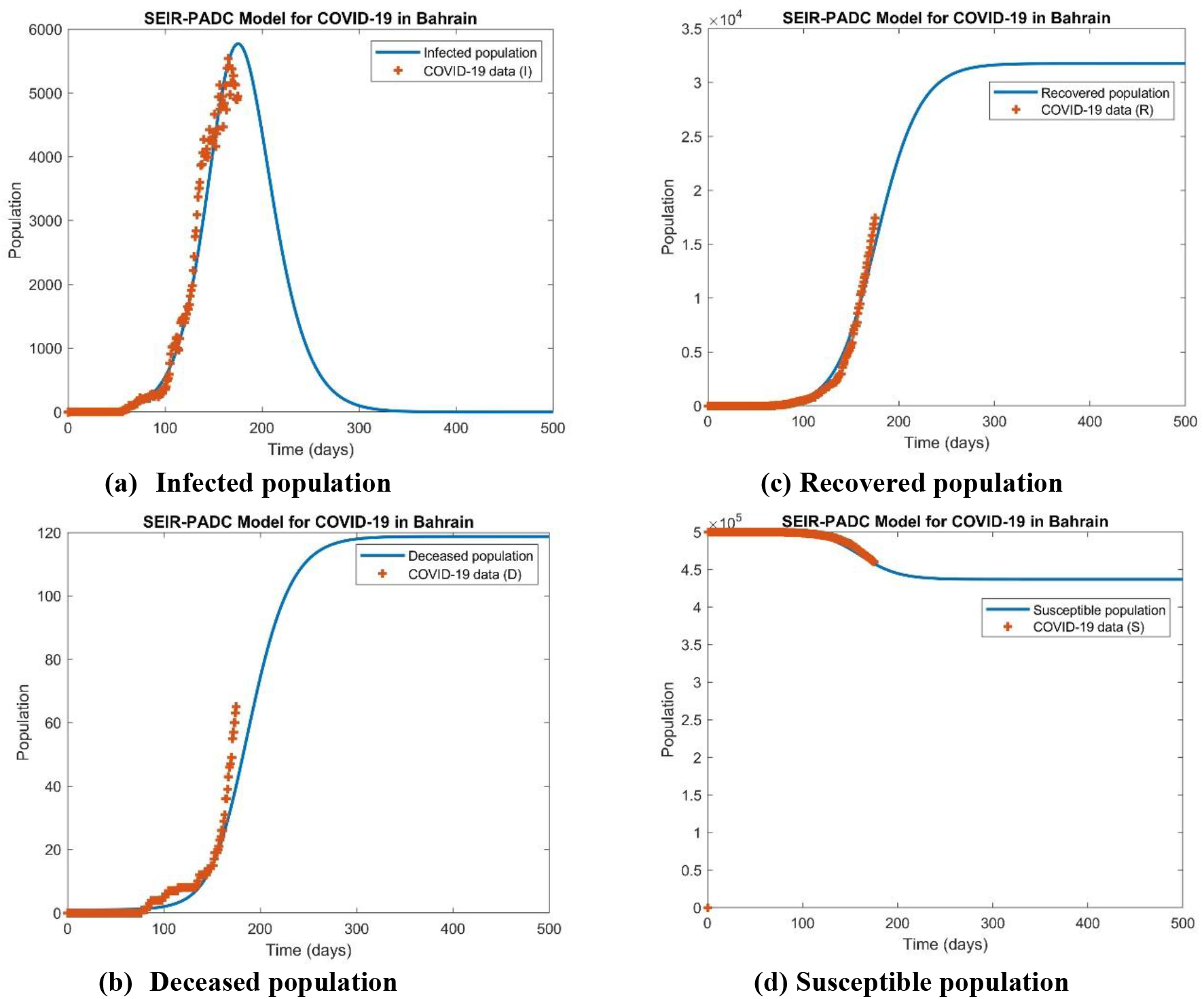
Fitting 4 set of COVID-19 data simultaneously with SEIR-PADC model for dynamics of COVID-19 in Bahrain (23 June 2020).

**Figure 4:**
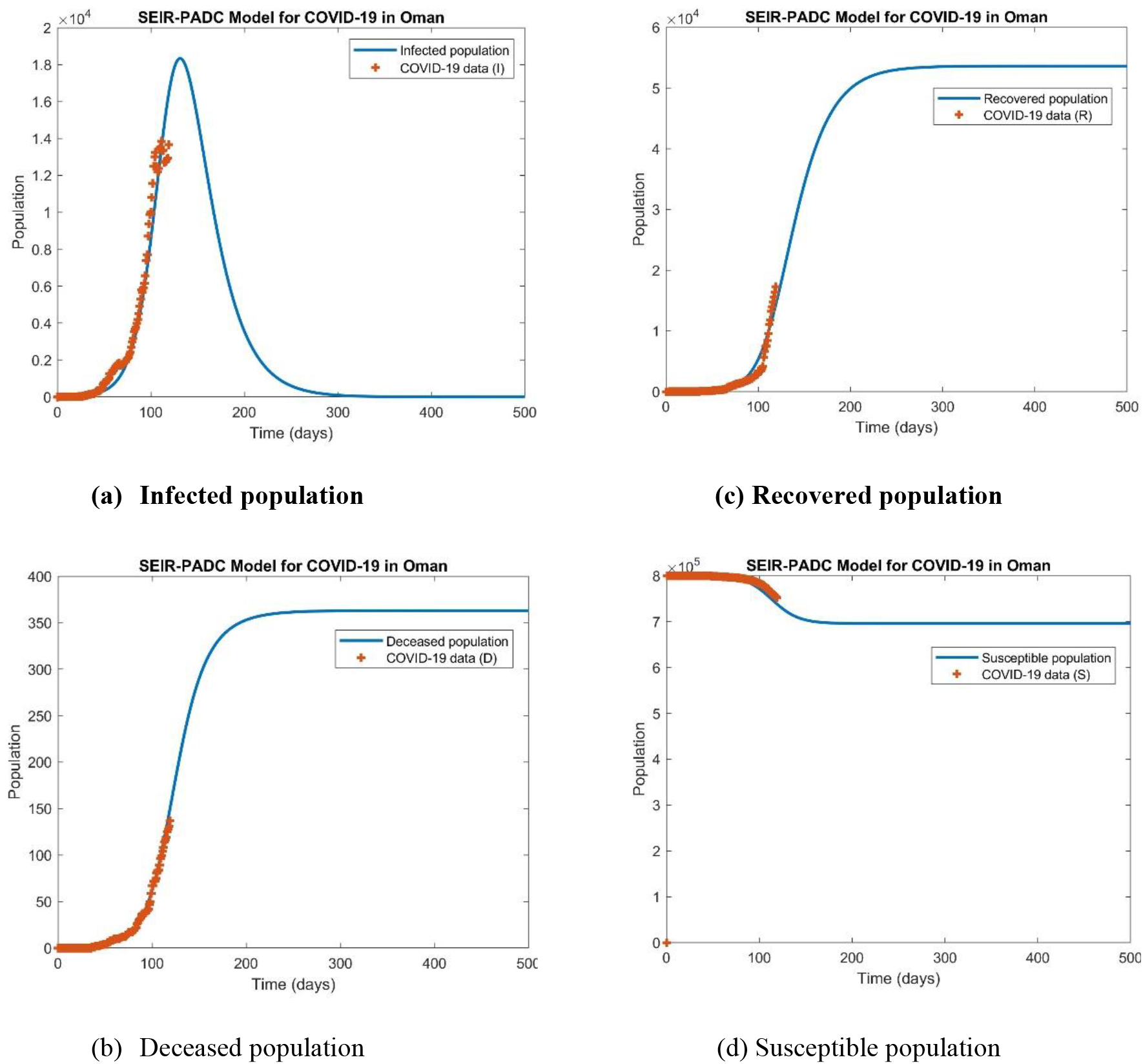
Fitting 4 set of COVID-19 data simultaneously with SEIR-PADC model for dynamics of COVID-19 in Oman (23 June 2020).

**Figure 5:**
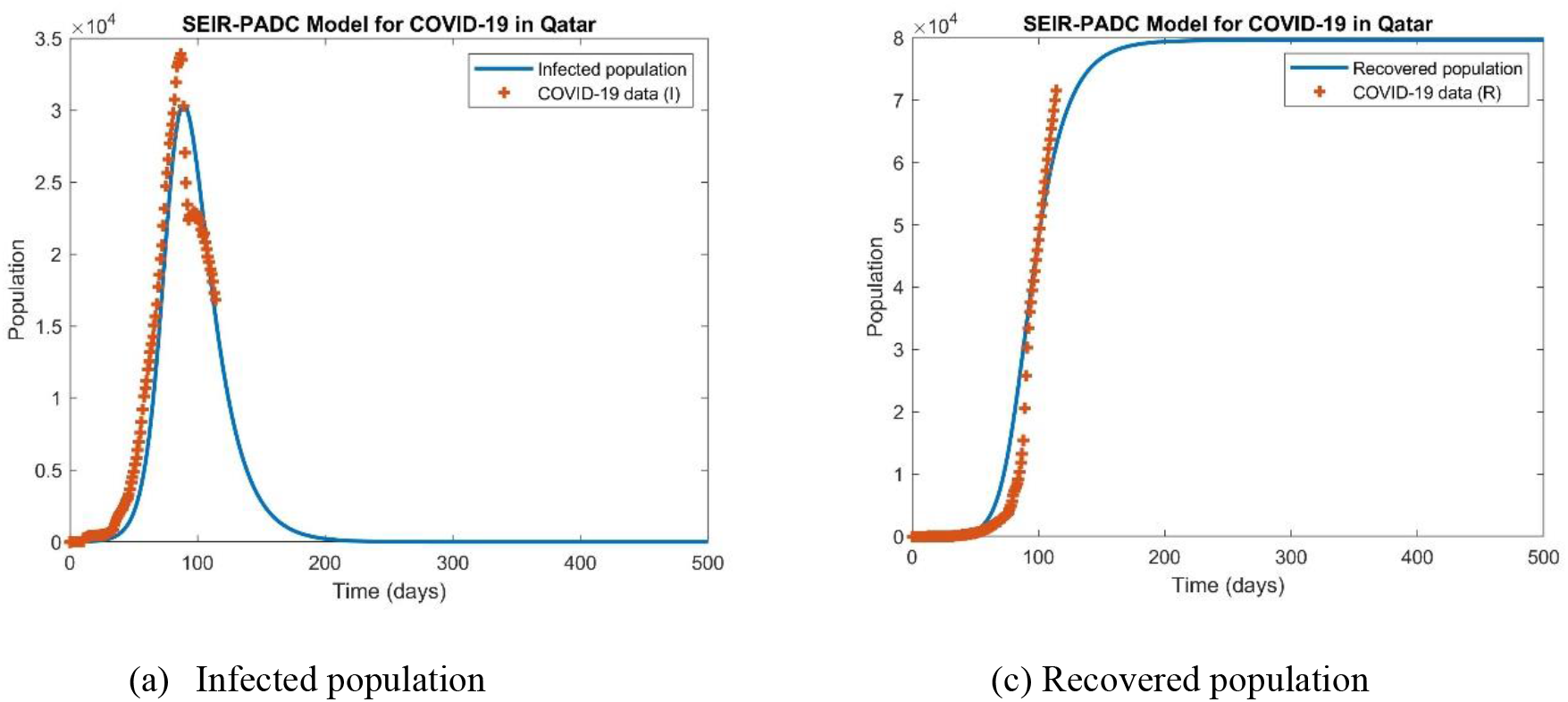
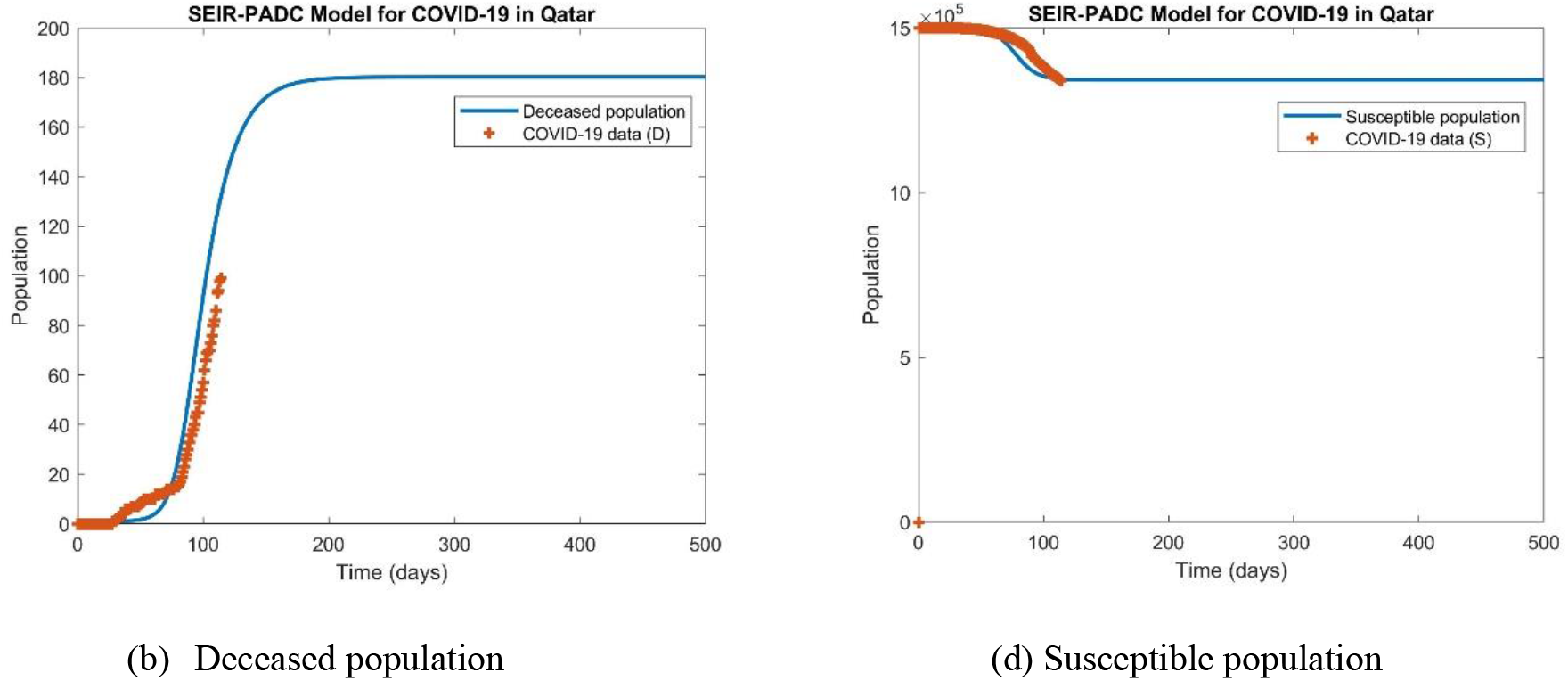
Fitting 4 set of COVID-19 data simultaneously with SEIR-PADC model for dynamics of COVID-19 in Qatar (23 June 2020)

**Figure 6:**
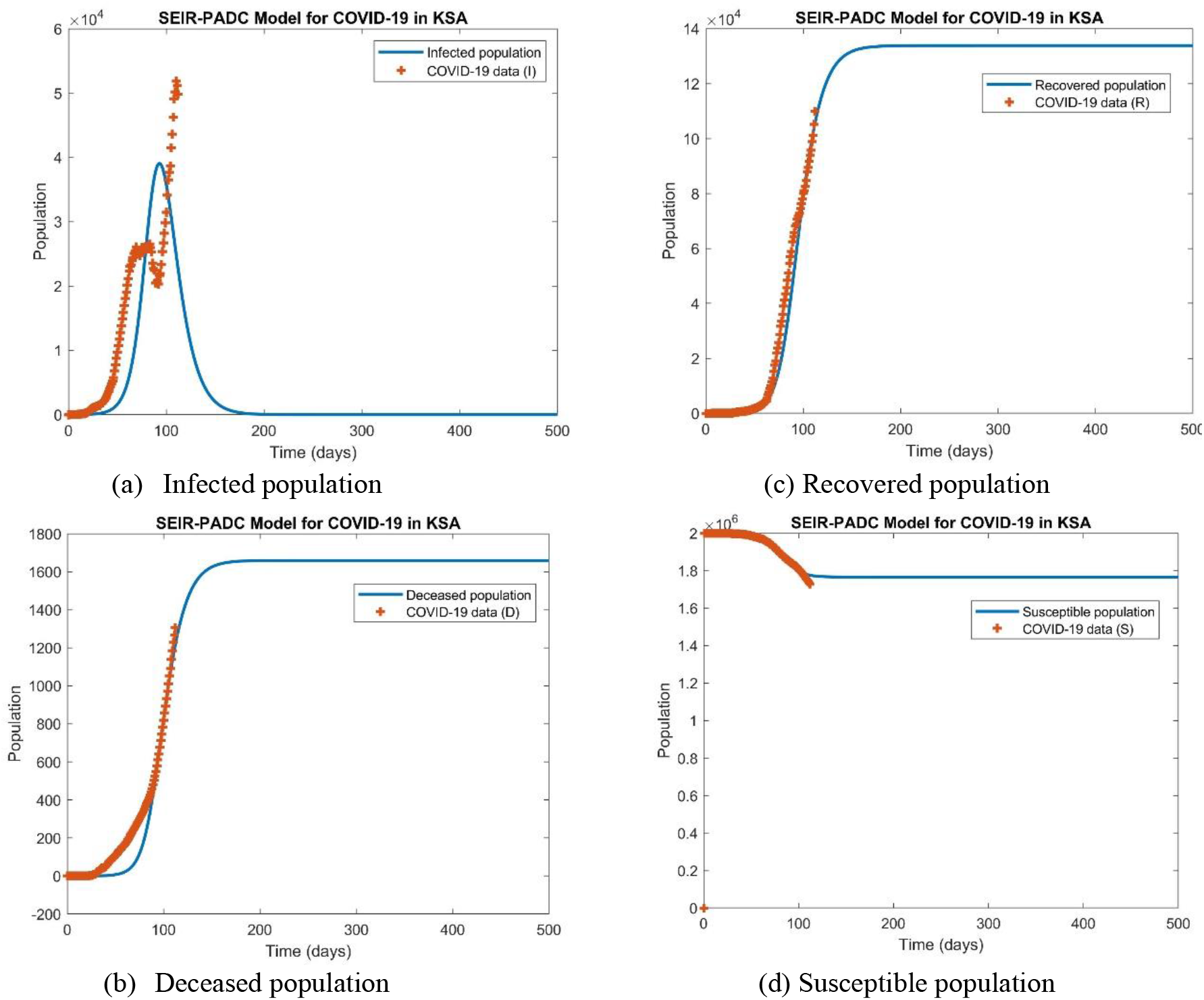
Fitting 4 set of COVID-19 data simultaneously with SEIR-PADC model for dynamics of COVID-19 in KSA (23 June 2020).

**Figure 7:**
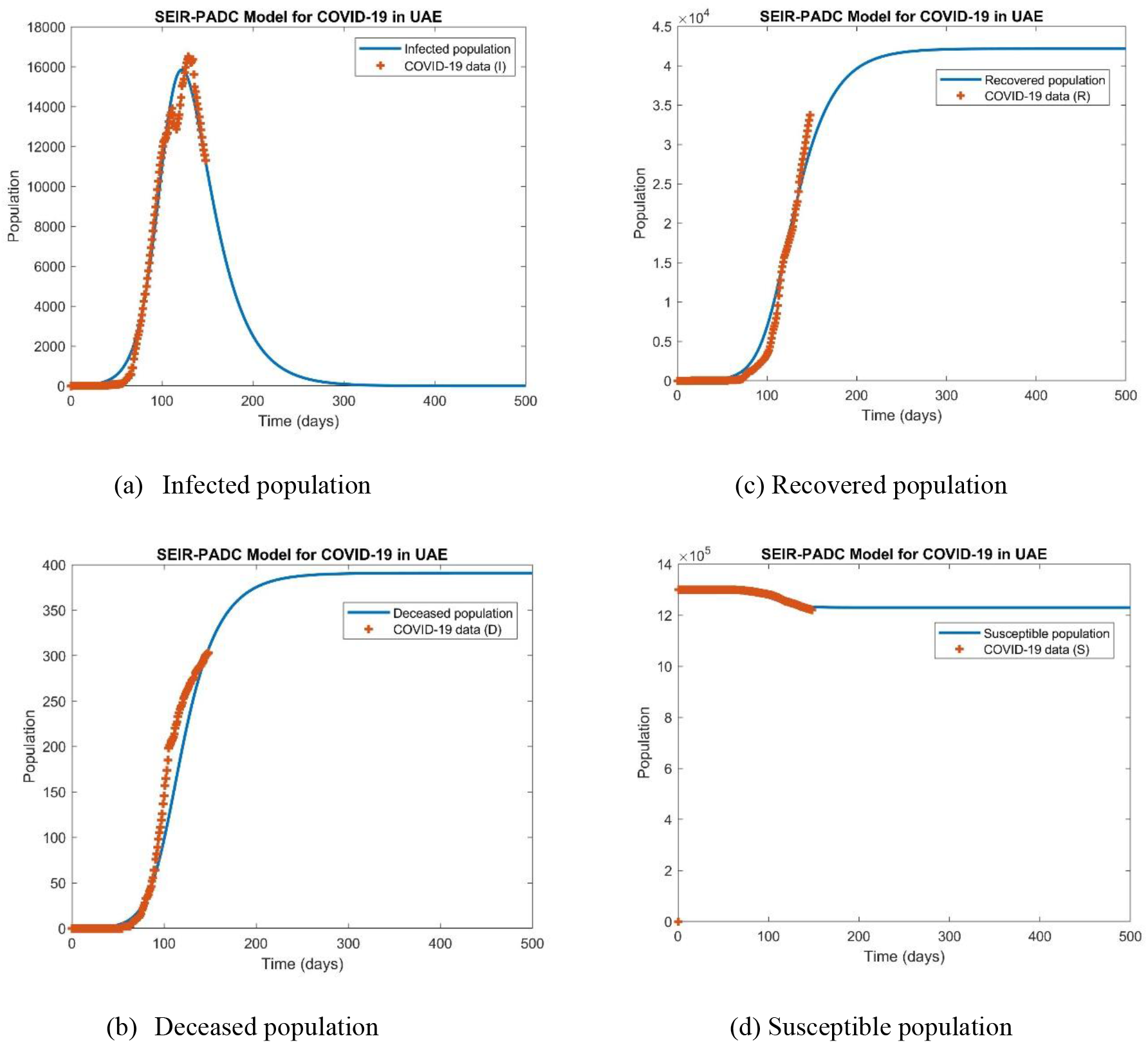
Fitting 4 set of COVID-19 data simultaneously with SEIR-PADC model for dynamics of COVID-19 in UAE (23 June 2020).

Important dates including peak number and day of infectious, peak value and day of people in critical condition, maximum recovered and deceased populations, and end of pandemic in Kuwait can be feasibly obtained from the Figure 2 and computed values from SEIR-PADC model.

Using the optimization algorithm (fminsearch) in MATLAB and initial conditions given in equations (10-11), the following values are found for the model coefficients (See TABLE 1).

**TABLE 1.**
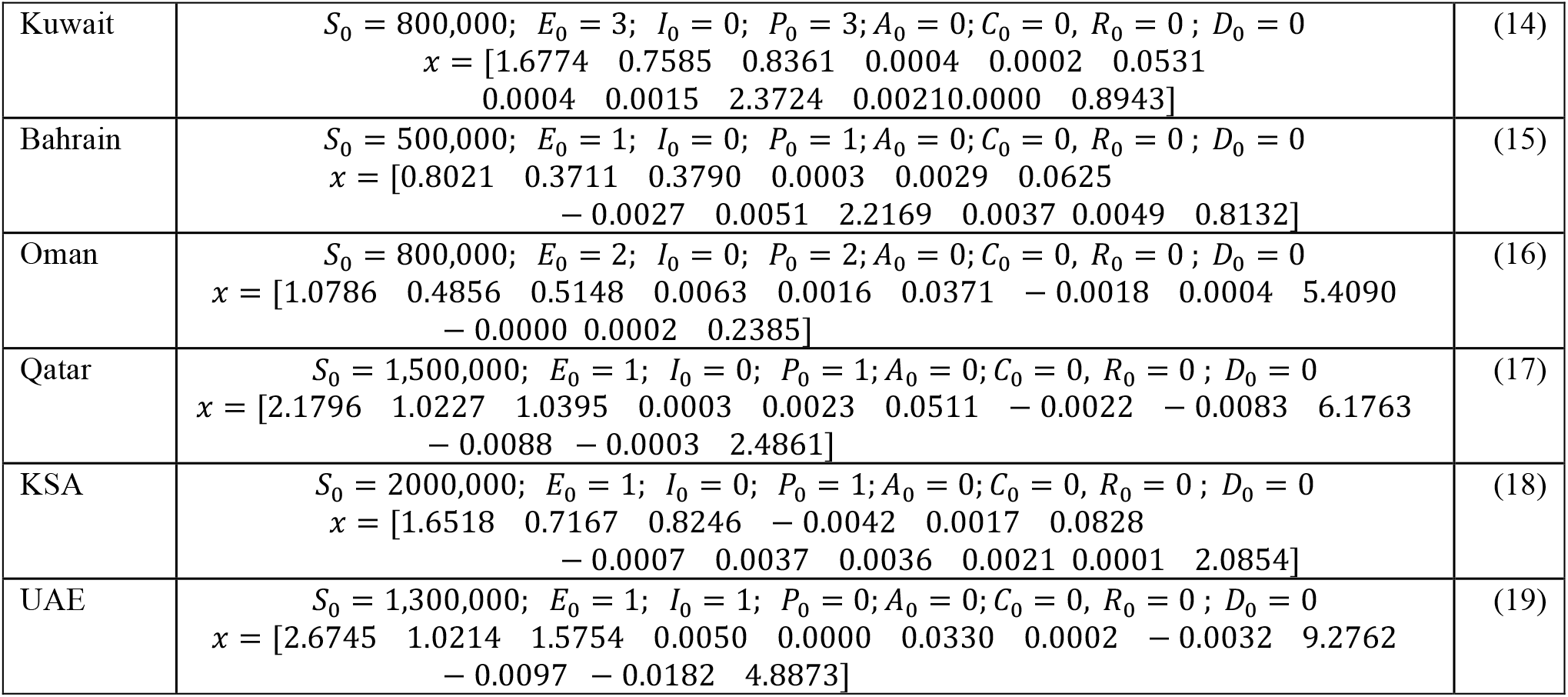
SEIR-PADC models’ results for individual countries

## IV. conclutions

The present research developed SEIR-PADC dynamic models for COVID-19 outbreak in GCC countries, i.e., Kuwait, Bahrain, Oman, Qatar, KSA, and UAE. The aim was to computationally fit simultaneously any available set of COVID-19 data for better prediction of trends of COVID-19 dynamics. From the results of this study, it can be concluded that. SEIR-PADC dynamic model was successfully implemented. Initial conditions for 12 unknown coefficients in SEIR-PADC model were correctly identified. MATLAB optimization with a convergence criterion is successfully applied to find best fitted prediction from SEIR-PADC model to COVID-19 data. Important peaks of population sizes and dates are predicted successfully indicting that Kuwait has passed all important peaks of critical hospitalization and peak of infectious populations. SEIR-PADC model is a simple and robust model to correctly pick up various well-defined population in SIR type models. The model has successfully fitted 5-set of COVD-19 data.

## Data Availability

data is public.

https://www.worldometers.info/coronavirus/

